# Development and qualification of an LC-MS/MS method for the quantification of MUC5AC and MUC5B mucins in spontaneous human sputum

**DOI:** 10.1101/2024.10.13.24315044

**Authors:** Weiwen Sun, Si Mou, Catherine Huntington, Helen Killick, Ian Christopher Scott, Aoife Kelly, Monica Gavala, Jessica Holmen Larsson, Deepika Vakkalanka, Neil E Alexis, Walter Wiley, Aaron Wheeler, Kumar Shah, Moucun Yuan, William R. Mylott, Kévin Contrepois, Anton I. Rosenbaum

**Affiliations:** Integrated Bioanalysis, Clinical Pharmacology and Safety Sciences, R&D, AstraZeneca, South San Francisco, CA 94080, USA; Biologics Engineering, R&D, AstraZeneca, Cambridge, UK; Translational Science and Experimental Medicine, Research and Early Development Respiratory & Immunology, R&D, AstraZeneca, Cambridge, UK; Translational Science and Experimental Medicine, Research and Early Development Respiratory & Immunology, R&D, AstraZeneca, Gaithersburg, MA 20878, USA; Chromatographic Services – Research & Development Biologics by LC–MS/MS, PPD Laboratory Services (a part of Thermo Fisher Scientific), Richmond, VA 23229, USA; University of North Carolina at Chapel Hill, Center for Environmental Medicine Asthma and Lung Biology, Chapel Hill, NC 27599, USA

**Keywords:** MUC5AC, MUC5B, LC-MS/MS, bioanalytical method, sputum collection, sputum processing, method qualification

## Abstract

**Aim:** Airway mucins are promising biomarkers in respiratory diseases. In this study, we aimed to identify a suitable sputum collection and processing method, as well as qualify a bioanalytical method for soluble MUC5AC and MUC5B quantification in clinical samples.

**Method:** Mucins were quantified in induced and spontaneous sputum collected from the same COPD patients and following various sample processing procedures. Our LC-MS/MS method used truncated recombinant mucins as surrogate analytes and a surrogate matrix approach.

**Results:** Frozen spontaneous sputum was found to be a suitable and convenient matrix for mucin quantification and fit-for-purpose method qualification was performed.

**Conclusion:** Our methodology provides accurate and reliable MUC5AC and MUC5B quantification and facilitates multi-site clinical studies in COPD and potentially other respiratory diseases.

## Introduction

Mucociliary clearance (MCC) is critical to maintain the health and function of the respiratory system — by acting as a physical barrier, maintaining hydration status, and removing inhaled environmental insults including microorganisms, aeroallergens, and pollutants such as smoke [1]. The major components of mucus are large, polymeric mucin glycoprotein macromolecules (2-50 MDa), for which up to 90% of the weight consists of glycans, that are critical for local host defence and MCC [2]. Of the thirteen mucins expressed in airways, seven predominate including two secreted gel-forming mucins (MUC5AC and MUC5B), four membrane mucins (MUC1, -4, -16, -20), and one secreted non-gel forming mucin (MUC7). MUC5AC and MUC5B are predominantly secreted by goblet cells and submucosal glands in the airway epithelium, respectively, and provide the biophysical viscoelastic properties required for airway mucus transport.

In respiratory diseases including chronic obstructive pulmonary disease (COPD) [3], cystic fibrosis [4], non-cystic fibrosis bronchiectasis (NCFBE) [5], and asthma [6], hyper-secretion of dysfunctional airway mucus is observed and associates with increased mucus plugging and airway obstruction, particularly in the distal airways. Thus, therapeutic interventions that reduce mucus hyper-secretion and restore healthy mucus composition, hydration or clearance are being investigated in patients diagnosed with respiratory diseases such as COPD [7].

Mucus plugging imaging through computed tomography (CT) scans assess the presence, extent, and distribution of mucus plugs which can aid in diagnosis, phenotyping, and treatment monitoring of lung conditions such as asthma and COPD [8-10]. While this technology has proven valuable for assessing a range of respiratory conditions, it is semi-quantitative and time-consuming which potentially limits its application in the clinic. Recent studies have shown that increased levels of soluble, secreted airway MUC5AC and 5B and altered MUC5AC/5B ratio associate with disease progression, lung function decline, cough, risk of exacerbations, and overall mortality of patients with chronic bronchitis [11,12]. Given their potential value as diagnostic, prognostic, and pharmacodynamic biomarkers in COPD and other respiratory diseases, there is a growing interest in developing accurate and reliable methodologies for quantification of soluble mucins.

However, mucin quantification in sputum (expectorated airway mucus) has proven challenging because of the heterogenous nature of the matrix and mucin physicochemical properties, including their extremely large size and heterogenous post-translational modification (PTM) profiles [2,13]. While methods such as enzyme-linked immunosorbent assay (ELISA) [14,15] and western blots [16,17] have been used to study mucins, they rely on antibodies whose binding affinity can be affected by different PTM profiles. In recent years, liquid chromatography–mass spectrometry (LC-MS) methods have been developed to quantify MUC5AC and MUC5B in lung-derived fluids (such as sputum and bronchoalveolar lavage fluid [BALF]) [11,12,18-20]. Those methodologies provide high throughput, sensitive, and specific analysis of mucins, however, quantification most often relies on spiked-in stable isotope-labeled peptides, which do not account for variability associated with sample preparation (*i.e.* reduction, alkylation, digestion) and matrix interference.

To circumvent those limitations, we developed and qualified a LC-MS/MS method for quantification of MUC5AC and MUC5B in human sputum using a surrogate analyte (*i.e.* truncated recombinant mucins) and a surrogate matrix approach. In addition, we compared various sputum collection and processing workflows and propose to normalize mucin quantities to total peptide content. Altogether, our approach is accurate, robust, and sensitive and is suitable for MUC5AC and MUC5B quantification in clinical sputum samples.

## Materials and Methods

### Generic reagents

Milli-Q reagent water (resistivity >16 MΩ/cm with total organic carbon <100 ppb) was produced in-house with a Milli-Q® IQ 7000 Ultrapure Lab Water System (Millipore, Burlington, MA, USA). Pierce™ protease inhibitor mini-tablets, Bond-Breaker™ TCEP solution (Neutral pH (0.5 M)), and Pierce™ quantitative colorimetric peptide assay kits were purchased from Thermo Fisher Scientific, Waltham, MA, USA. Pierce™ MS grade trypsin protease was purchased from Thermo Fisher Scientific, Waltham, MA, USA and VWR International, Radnor, PA, USA. 10x Dulbecco’s phosphate buffered saline (PBS) was purchased from Mediatech Inc., Manassas, VA, USA. Fluka^TM^ formic acid (∼98%) and 1x Dulbecco’s PBS were purchased from Sigma, St. Louis, MO, USA. Bovine serum albumin (BSA), 2-chloroacetamide, CHROMASOLV Plus 2-propanol (isopropanol; IPA) for HPLC (99.9%), and CHROMASOLV LC-MS acetonitrile (ACN) were purchased from Sigma-Aldrich, Burlington, MA, USA. Ammonium bicarbonate, ACN with formic acid, and water with formic acid were purchased from VWR International, Radnor, PA, USA. Amylase assay kit was purchased from Abcam, Boston, MA, USA.

### Critical reagents

Truncated recombinant human MUC5AC (rH-MUC5AC) and MUC5B (rH-MUC5B) were produced in Chinese hamster ovary (CHO) cells with an unglycosylated molecular weight of 266 kDa and 240 kDa, respectively (**Supplemental Figure 1**). Intact, unglycosylated MUC5AC and MUC5B molecular weights are 585 kDa and 596 kDa, respectively. Expression constructs were made of N-terminal domain-tandem repeats and C-terminal domain fragments flanked at N-terminus with Twin Strep tag and at C-terminus with His10 Tag for sequential affinity chromatography purification (HisTrap Excel and StrepTrap XT). Construct designs were inspired by previously described constructs for MUC5AC and MUC5B [21,22]. MUC5AC and MUC5B concentrations (2.360 mg/mL and 3.365 mg/mL) and identity (61% and 73%) were determined by amino acid and LC-MS/MS analyses, respectively. Out of six peptides tested, a single quantitative signature peptide which passed accuracy and precision (A&P) as well as matrix parallelism acceptance criteria was selected per mucin protein (**Supplemental Table 1**). Corresponding internal standards (IS) were obtained from 21^st^ Century Biochemicals Inc., Marlborough, MA, USA. IS stock solutions were prepared by dissolving neat reference standard material into the appropriate volume of solvent (water). Stock solutions were stored at -80°C.

### Patient recruitment and institutional review board consent

Assessment of sputum collection and processing workflows were performed using sputum from five patients diagnosed with COPD (following Global Initiative for Chronic Obstructive Lung Disease criteria [23] recruited for research studies at the Medicines Evaluation Unit [Manchester University NHS Foundation Trust, Manchester, UK]). This research was approved by local ethics committees (Manchester South, UK; REC reference 06/Q1403/156) and all patients provided written informed consent. Patient inclusion criteria consisted in the presence of cough and sputum on most days for ≥ 3 months/year in at least the 2-year period prior to the study.

Primary matrix quality controls (QCs) and individual lots used for specificity/selectivity and matrix parallelism assessment were from patients with moderate-to-severe COPD and chronic bronchitis. Patients were required to have a documented history of COPD or chronic bronchitis for at least 1 or 2 years, respectively. This study was approved by local independent ethics committees and institutional review boards and was carried out in accordance with the International Council for Harmonization Guidance for Good Clinical Practice. All patients provided written informed consent and human subject confidentiality and privacy were ensured by de-identification of all biological samples.

### Sputum collection

Both induced and spontaneous sputum were collected as previously described [24,25]. Briefly, patients’ pulmonary function was first tested by spirometry prior to being given 2-4 puffs of Ventolin to reduce the irritant effect of the salt solution on their airways. After approximately 20 min and post bronchodilator test was recorded (forced expiratory volume in 1 second [FEV1]), subjects inhaled (using tidal breathing) a hypertonic saline solution produced by a handheld nebulizer. Induced sputum was then coughed into a sterile specimen cup after patients blow their nose and rinse their mouth. This procedure was repeated three times and induced sputum samples were split into two aliquots, one of which was placed on ice (referred as “fresh”), and the other was snap-frozen and transferred to -80°C for storage (referred as “frozen”). Spontaneous sputum was produced naturally, without an inhaled stimulus, and collected into a sterile specimen cup. Spontaneous sputum samples were snap-frozen on dry ice and transferred to -80°C. Spontaneous sputum was used for primary matrix QCs as well as specificity/selectivity and matrix parallelism assessment.

### Sputum processing

Sputum samples were processed as previously described [26]. Briefly, fresh sputum samples were kept on ice while frozen sputum samples were thawed at room temperature prior to processing. Mucus plugs were selected from the sputum samples, weighed and solubilized with cold Dulbecco’s PBS at a ratio of 1:8 (w:v) by pipette actuating 8-10 times followed by homogenization on rotating tumbler for 15 min at room temperature. A PBS fraction was created by centrifugation at 790 rcf for 10 min at 4°C. A dithiothreitol (DTT) fraction was generated by solubilizing pellets with the same volume of 0.1% DTT following the same procedure. Both PBS and DTT fractions were aliquoted, snap-frozen, and stored at -80°C. Only DTT fractions were generated for samples used for primary matrix QCs as well as specificity/selectivity and matrix parallelism assessment.

### Amylase assay

2 – 50 μL of processed sputum samples (final volume of 50 μL/well with ddH2O) in duplicates were aliquoted for amylase analysis using an amylase assay kit (Abcam, Boston, MA, USA) following the manufacturer’s instructions. Wavelength absorption was measured at OD 405 nm with sensitivity > 0.2 mU/well.

### Sample preparation for LC-MS/MS analysis

Processed sputum samples were thawed on ice, vortexed and a 25-μL aliquot was transferred to a new 96 well plate (deepwell, LoBind) and diluted 2-fold with 25 μL of 1.00 mg/mL BSA in 1x PBS (surrogate matrix). A 50 μL aliquot of 1x PBS with protease inhibitor was added to each sample well. The plate was sealed, vortexed (800 rpm, 2 min), sonicated (5 min), incubated (10 min, 95°C), and allowed to cool. Under yellow light conditions, 22 μL of 62.5 pmol/mL IS working solution (prepared in 50 mM TCEP) was added to all wells, except for double blank wells (blanks without IS). For double blank wells, 22 μL of 50 mM TCEP was added. The plate was sealed, centrifuged (800 rpm, 1 min), vortexed (800 rpm, 2 min), incubated (10 min, 95°C), and allowed to cool. A 10-μL aliquot of 400 mM 2-chloracetamide solution was added to each well. Prior to incubation (30 min, 37°C) and cooling, the plate was again sealed, centrifuged, and vortexed (as above). Trypsin solution (10 μL of 0.2 mg/mL) was added to each sample. Prior to overnight incubation (∼16-20 h, 37°C), the plate was sealed, centrifuged, and moderately vortexed (as above). The following day, proteolytic digestion was quenched with 10 μL of 1.5% (v/v) formic acid in water. The plate was sealed, vortexed (800 rpm, 2 min), and centrifuged (3700 rcf, 10 min). A Sorenson liquid handling system was used to semi-automate the transfer of supernatant (100 μL per sample) to a new 96-well plate.

### LC-MS/MS data acquisition

Chromatographic separation was achieved using a reversed phase analytical column maintained at 50°C (ACQUITY UPLC BEH C18, 130Å, 2.1 mm X 50 mm, 1.7 µm, Product No. 18602350, Waters Corp., Milford, MA, USA) with 0.1% (v/v) formic acid in water (mobile phase A) and 0.1% (v/v) formic acid in acetonitrile (mobile phase B). Flow rate was constant at 0.6 mL/min, total run time was 8.0 min and injection volume was 25 μL. Mobile phase gradient is described in **Supplemental Figure 2**. Samples were analysed using a 6500+ triple quadrupole mass spectrometer (SCIEX, Framingham, MA, USA) operated in positive electrospray ionization mode and instrument parameters can be found in **Supplemental Table 2**.

### Quantitative colorimetric peptide assay

20 μL sample volume was transferred to a 96-well Nunc Maxisorp plate (Thermo Scientific) for total peptide analysis using Pierce™ quantitative colorimetric peptide assay kit following the manufacturer’s instructions. Wavelength absorption was measured on a Tecan SPARK Plate Reader (OD 480 nm).

### Calibration standards preparation

Due to the endogenous nature of mucins in human sputum, a solution of 1.00 mg/mL BSA in 1x PBS served as an analyte-free surrogate matrix. Calibration standards (CALs) were prepared in surrogate matrix with rH-MUC5AC concentrations of 0.125, 0.250, 0.500, 2.00, 5.00, 20.0, 80.0, and 100 µg/mL and rH-MUC5B concentrations of 0.1875, 0.375, 0.750, 3.00, 7.50, 30.0, 120, and 150 µg/mL, respectively. During method qualification, CALs were freshly prepared (in polypropylene tubes and never frozen) for each run.

### Quality control preparation

Surrogate matrix quality controls (QCs) were prepared in 1.00 mg/mL BSA in 1x PBS with rH-MUC5AC concentrations of 0.125 (LLOQ), 0.375 (Low), 37.5 (Mid), and 75.0 µg/mL (High), and rH-MUC5B concentrations of 0.1875 (LLOQ), 0.563 (Low), 56.3 (Mid), and 113 µg/mL (High). The lower limit of quantitation (LLOQ) was defined as the lowest non-zero concentration that could be quantitatively determined with acceptable accuracy and precision (see details below). Surrogate matrix QCs were prepared in polypropylene tubes, thoroughly mixed, and sub-aliquoted into daily use portions, which were stored at - 80°C. At least one accuracy and precision run, and all matrix stability runs, used freshly prepared (never frozen) surrogate matrix QCs.

Due to relatively high endogenous analyte concentrations from available sputum lots, assessment of true, low-level primary matrix QCs was not feasible. As such, unfortified primary matrix from 11 COPD patients served as the endogenous-QCs (mid-level). Overspike-QCs were prepared by spiking endogenous-QC pools with additional analyte: 50.0 µg/mL of rH-MUC5AC or 10.0 µg/mL of rH-MUC5B. For MUC5AC, the resultant primary matrix endogenous- and overspike-QC nominal concentrations were established as 18.2 and 62.4 µg/mL, respectively. For MUC5B, the resultant primary matrix endogenous- and overspike-QC nominal concentrations were established as 80.2 and 81.6 µg/mL, respectively. Theoretical concentrations for these primary matrix QCs were based on mean concentration values from the core precision and accuracy runs (inter-assay). Additional over-the-curve QCs (OTC-QCs) were prepared by spiking pooled primary matrix with rH-MUC5AC or rH-MUC5B such that OTC-QC concentrations for MUC5AC and MUC5B were 218 μg/mL and 230 μg/mL, respectively. Primary matrix QC pools were prepared in polypropylene, thoroughly mixed, and daily use portions were stored at -80°C. Owing to the endogenous nature of the analytes, frozen primary matrix QCs were used throughout the evaluation.

### Method qualification studies

Fit-for-purpose method qualification studies included assay linearity, accuracy and precision, specificity/selectivity, analyte stability, and matrix parallelism. Accuracy and precision were calculated as percent difference from theoretical (%DT) and percent coefficient of variation (%CV):

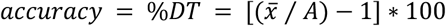

and

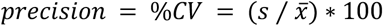

where 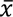 is the calculated mean concentration, *A* is the theoretical concentration, and *s* is the calculated standard deviation. Data from the core accuracy and precision runs were used to establish endogenous concentrations for primary matrix QCs and for lots of primary matrix; in turn, these endogenous concentrations were used during the execution of subsequent stability and dilutional linearity studies. During the matrix parallelism studies, mean intra-dilutional concentration was corrected for each dilution factor. Intra-lot precision was calculated as follows:

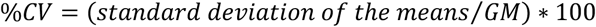

where *GM* is the grand mean across all dilutions of a human sputum lot.

### Acceptance criteria

Unless otherwise specified, acceptance criteria for qualification studies were ≤25%CV and ±25%DT at LLOQ, and ≤20.0%CV and ±20.0%DT for all other (non-LLOQ) levels. To demonstrate matrix parallelism, 5 out of 6 human sputum lots must exhibit intra-lot precision ≤20.0% CV.

### Data analysis and visualization

LC-MS/MS data were acquired and processed in Analyst (v1.6.3, SCIEX), MultiQuant (v3.0.3, SCIEX) using the MQ4 processing algorithm, and Assist Laboratory Information Management System (Assist LIMS) software (PPD, Wilmington, NC, USA). Summary statistics from qualification studies were calculated in Assist LIMS. All graphical displays featured in this manuscript were prepared in Prism v8.3.1 for Windows (GraphPad Software, San Diego, CA, USA).

## Results and Discussion

Soluble airway mucins, and in particular MUC5AC and MUC5B, are emerging as potential diagnostic, prognostic and pharmacodynamic biomarkers in COPD and other respiratory diseases. In this context, we compared various sputum collection and processing methodologies and qualified LC-MS/MS assays for MUC5AC and MUC5B quantification in clinical samples using surrogate analytes and a surrogate matrix approach.

### LC-MS/MS method development

To deliver accurate and reliable mucin concentrations in sputum samples, we designed a methodology using surrogate analytes (*i.e.* truncated recombinant human MUC5AC and MUC5B proteins) which were produced in a mammalian cellular system to harbour a complex glycosylation pattern and account for the variability associated with sample preparation and matrix interference. To our knowledge, this is the first attempt to use recombinant human MUC5AC and MUC5B protein standards for endogenous mucin quantification. Given the unavailability of mucin-free sputum, we employed a surrogate matrix approach to establish calibration standards and quality controls. The appropriateness and acceptability of the surrogate analytes and surrogate matrix approach were demonstrated with accuracy and precision (A&P) and matrix parallelism evaluations, respectively, during method qualification. In an effort to simplify sample preparation protocol and maximize assay throughput, mucins were denatured with heat instead of a denaturing agent such as guanidine hydrochloride or trifluoroethanol [11,12,18,19]. Therefore, samples could be directly analysed by LC-MS/MS without requiring prior sample clean-up (**Figure 1**).

**Figure 1.**
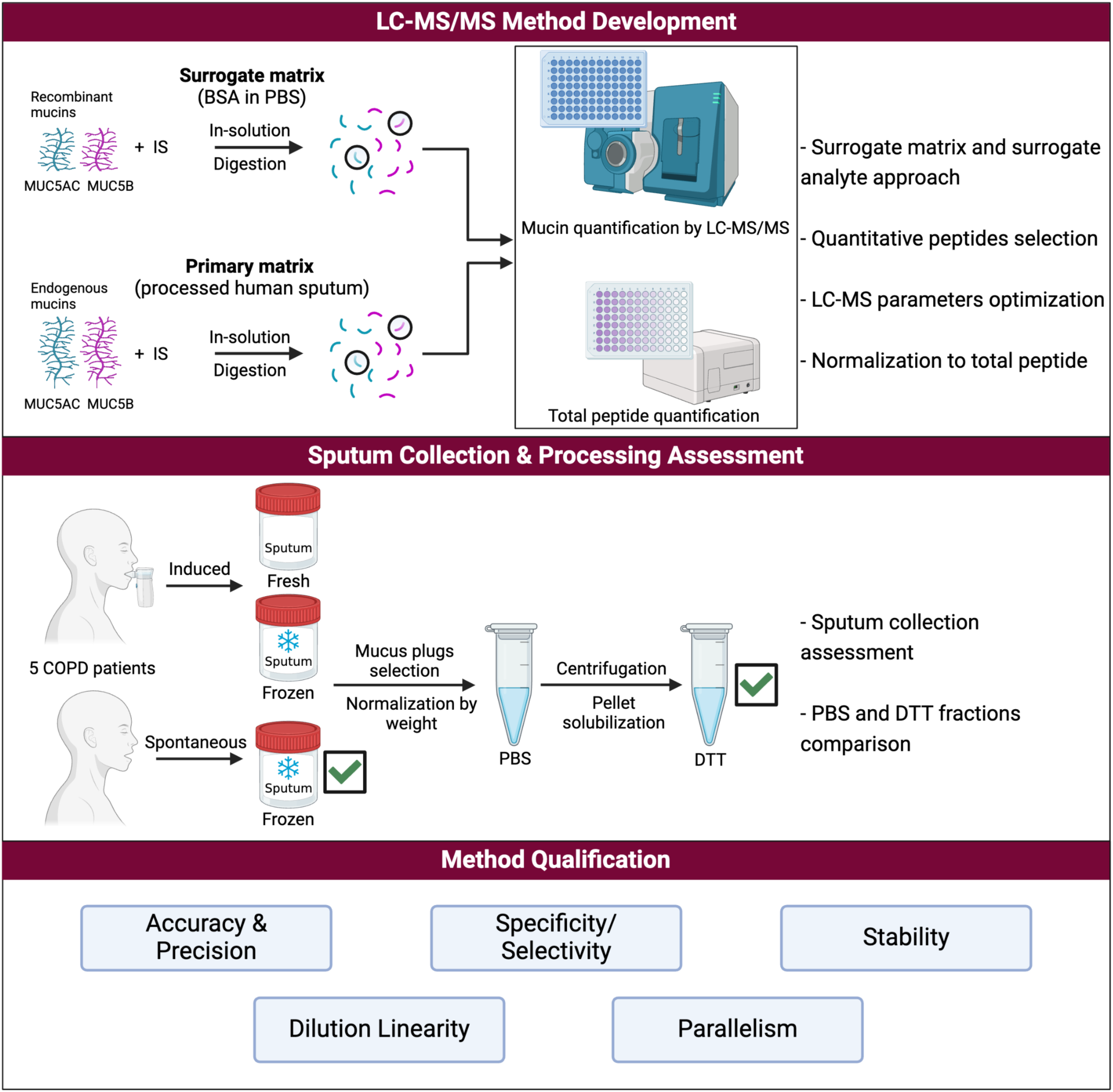
Summary of the LC-MS/MS method development, sputum collection and processing assessment, and method qualification. IS: internal standard; BSA: bovine serum albumin; PBS: phosphate buffered saline; DTT: dithiothreitol. Created with BioRender.com.

**Table 1.**
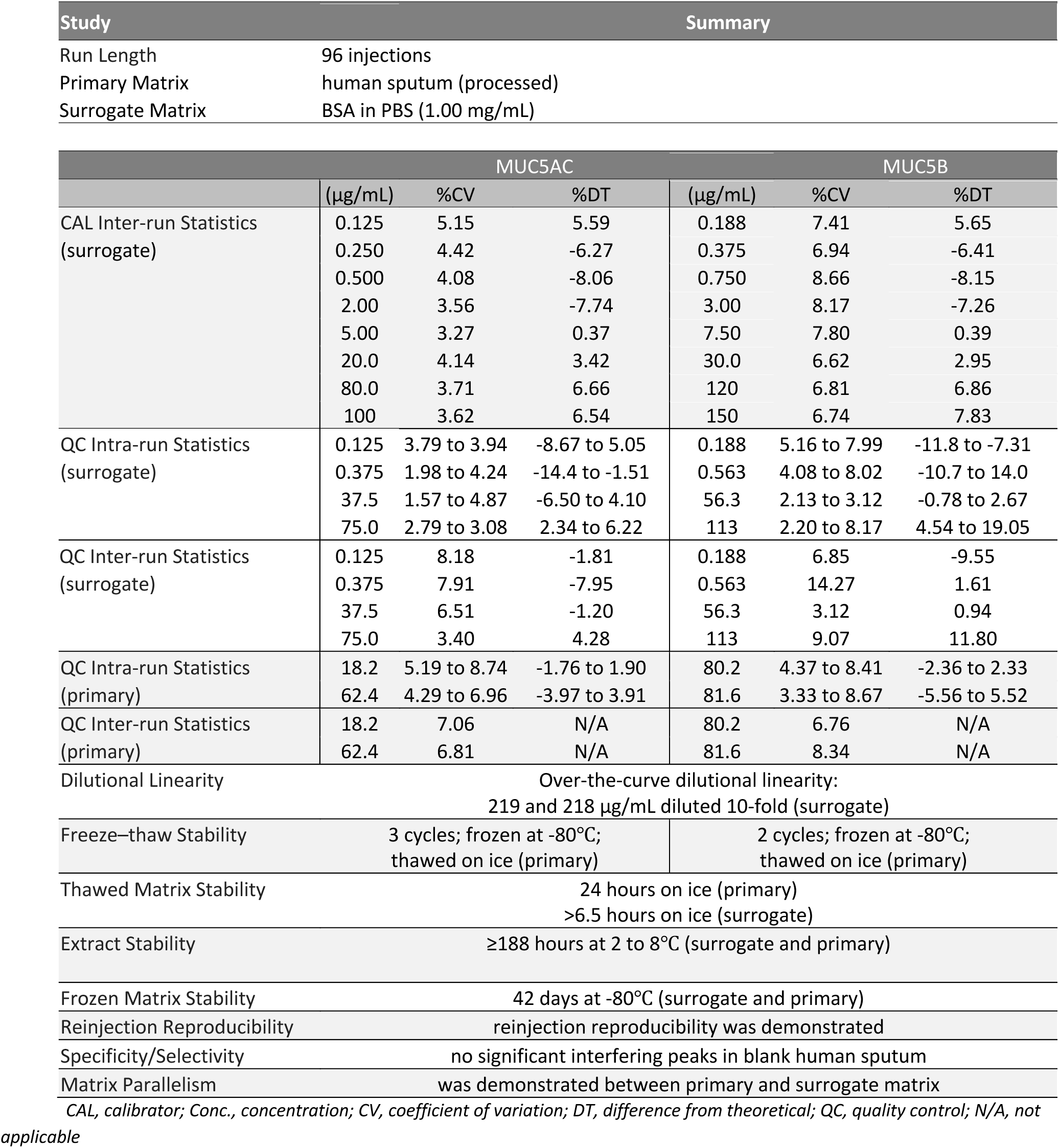
Summary of method qualification study results.

A significant challenge in mucin quantification is the selection of reliable proteotypic peptides. Indeed, it has been reported that quantification of mucins can be highly variable when using different peptides [20]. To overcome this limitation, several studies used the average or median concentrations of multiple peptides (n = 3-5) for mucins quantification [11,19,20], while others have used a single peptide [18]. We investigated six commonly detected MUC5AC and MUC5B tryptic peptides [11,19] and selected a single peptide per protein which passed A&P as well as matrix parallelism acceptance criteria (GTFLDDTGK for MUC5AC, and TWLVPDSR for MUC5B). Importantly, these peptides are located outside of tandem repeat (VNTR) domains rich in serine, threonine and proline (PTS) which are the primary sites of O-linked mucin-type glycosylation (**Supplemental Figure 1**). Following signature quantitative peptide selection, reverse phase chromatographic separation and quantitative multiple reaction monitoring (MRM) parameters were optimized for suitable peak retention time and enhanced peak intensity (see Material and Methods section).

### Sputum collection and processing assessment

Induced and spontaneous sputum collection are two frequently used methods for acquisition of sputum samples in the clinics [27,28]. Sputum induction has been historically the preferred procedure because it enables consistent sampling using a standardized process, produces higher sample yield, and is especially useful for patients who cannot produce sputum spontaneously [29-31]. However, it is more technically demanding and can cause patient discomfort. On the other hand, spontaneous sputum collection is simpler and more practical, but may suffer from inconsistent sample yield and potential contamination from saliva [32]. After collection, sputum samples are typically processed and then frozen prior to analysis.

Since it is unclear how various sputum collection procedures and processing workflows impact mucins quantification, we evaluated two types of sputum collection protocols, the effect of freezing sputum prior to downstream processing (*i.e.* fresh induced sputum, frozen induced sputum, and frozen spontaneous sputum) and two processed fractions namely PBS and DTT from five COPD patients (**Figure 1**, see Materials and Methods section). Patients’ demographics information can be found in **Supplementary Table 3**. MUC5AC and MUC5B concentrations were determined using the optimized LC-MS/MS method described above. Consistent with previous studies, MUC5AC concentrations were lower and distributed across a larger range than MUC5B [11,12] (**Figure 2**). In addition, we observed less mucins in PBS fractions in comparison to DTT fractions presumably because a reducing agent helps mucin solubilization. Thus, DTT fraction was selected as the most appropriate fraction for downstream analysis.

**Figure 2.**
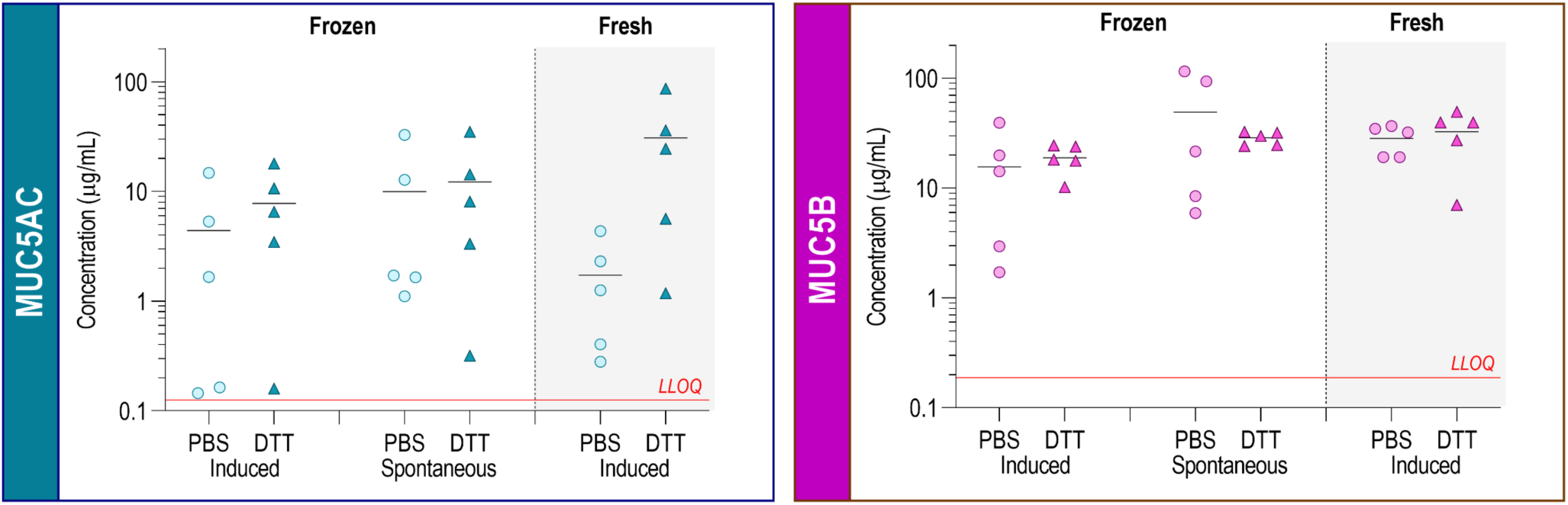
Univariate dot plots comparing different sample collection and processing methods. Each data point represents the mean of measurements from duplicate samples from 5 patients diagnosed with COPD. Concentration values (μg/mL) are plotted against the y-axis. Horizontal black lines = mean concentration; horizontal red lines = LLOQ, lower limit of quantification. PBS: phosphate buffered saline; DTT: dithiothreitol.

Next, we investigated how well MUC5AC and MUC5B concentrations in DTT fractions correlated across the different sputum collection and processing procedures. While MUC5AC levels correlated well across all the conditions with coefficients of determination >0.90, MUC5B concentrations in different collection methods were not well correlated (**Figure 3**). This difference may be explained by a narrower MUC5B concentration range and a greater impact of sputum collection protocols. We hypothesized that normalizing mucin concentrations to total peptide amount may provide a more reliable measurement across sputum conditions. Indeed, even with a limited number of specimens, we observed that correlations were improved after normalization with a coefficient of determination of 0.58 for MUC5B (**Figure 3**). Spontaneous sputum collection is susceptible to saliva contamination and saliva contains MUC5B. Saliva contamination was assessed using an amylase assay and was deemed negligible with low levels of amylase activity (i.e. 0.6-1.6%) in comparison to levels found in normal human saliva [33]. Altogether, we demonstrate that frozen spontaneous sputum is a suitable and practical matrix for mucin quantification after normalization to total peptide which facilitates mucin analysis in large multi-site clinical studies.

**Figure 3.**
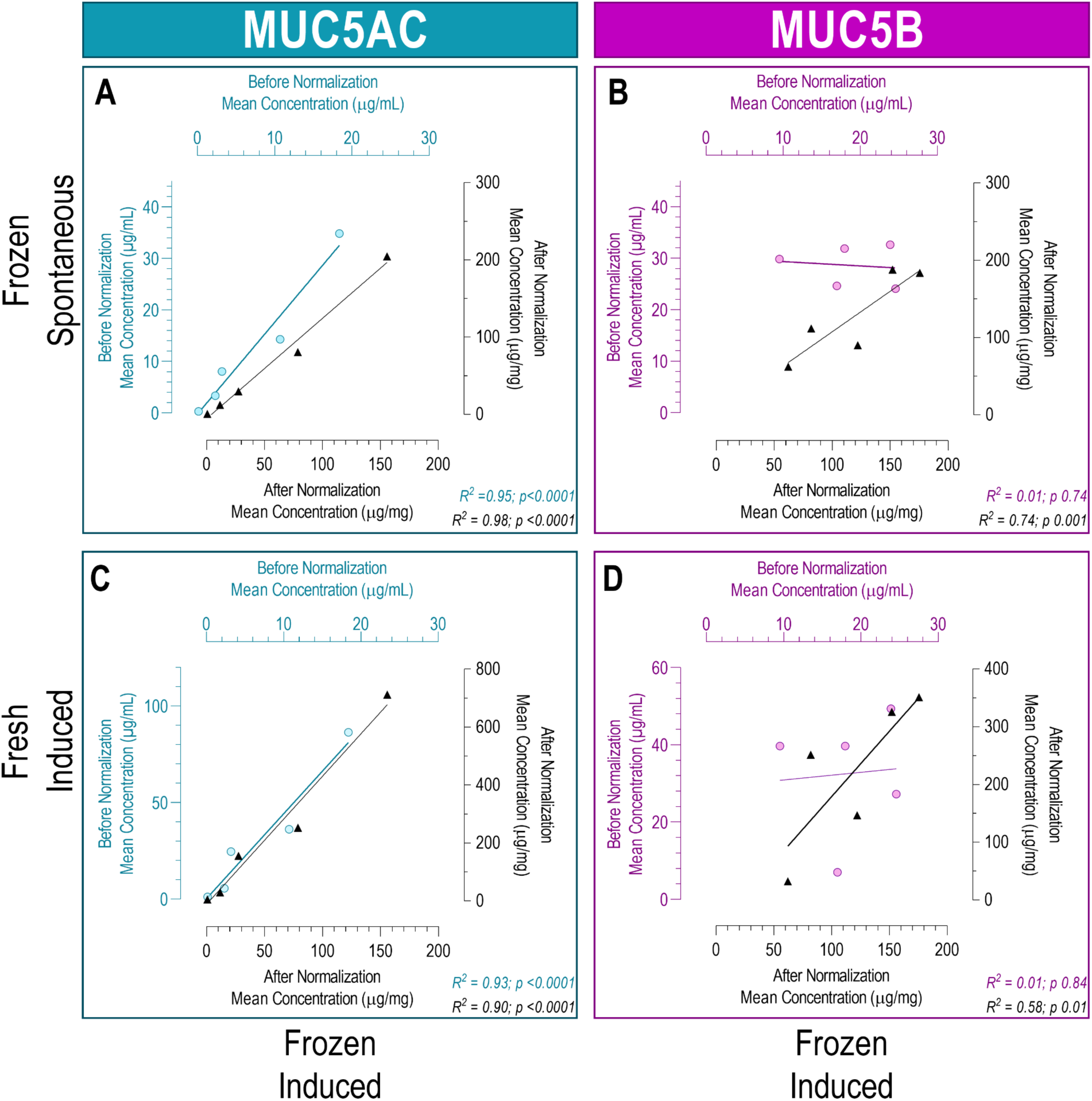
Scatter plots comparing analyte concentrations in DTT fractions from fresh induced, frozen induced and frozen spontaneous sputum before (colored axes) and after total peptide normalization (black axes) for MUC5AC (A-C) and MUC5B (B-D). Each data point represents the mean of measurements from duplicate samples. Unweighted linear regression lines, R^2^ and p-values are shown for each data set.

### Method qualification

Next, the LC-MS/MS method was qualified including run length evaluation, assay linearity, accuracy and precision, specificity/selectivity, analyte stability, dilutional linearity, as well as matrix parallelism.

#### Run length evaluation

To simulate the expected batch size and duration of an analytical run, surrogate matrix blanks were extracted and analysed in a single analytical run (one 96 well plate, 96 injections) with CALs and QCs distributed throughout the injection sequence. CAL and QC results indicated consistent signal response for both the analytes and the internal standards across the length of the run. Therefore, this 96-injection run length evaluation was deemed acceptable with no evidence of system or assay performance degradation during the analytical run.

#### Linearity, accuracy, and precision

Two eight-point calibration curves were included in each analytical run. A 1/concentration² weighted, least-squares regression model was used to evaluate the relationship between peak area response ratio (analyte response:internal standard response) and theoretical concentration. Best fit regression models were used to determine empirical concentrations and representative calibration curves and chromatograms are presented in **Supplemental Figure 3** and **Supplemental Figure 4**.

The accuracy and precision (A&P) evaluation comprised two analytical runs that spanned multiple days. These A&P runs included surrogate matrix QCs, which were prepared at LLOQ, low-, mid- and high-QC levels. Relatively high endogenous analyte concentrations from available sputum lots precluded the preparation and inclusion of low-level primary QCs so primary matrix QCs were only prepared at mid- and high -QC levels. Neat (unfortified) primary matrix pools served as mid-level QCs (also referred to as endogenous-QCs) and high-level QCs (also referred to as overspike-QCs) were created by over-spiking mid-level QCs with recombinant mucins as described in the Material and Methods section.

For MUC5AC, intra and inter-day accuracy and precision in surrogate matrix ranged from -14.4 to 6.22 %DT with %CV ≤ 8.18. In primary matrix the %DT range was from -3.97 to 3.91, with %CV ≤ 8.74 (**Figure 4B and C**). For MUC5B, intra and inter-day accuracy and precision in surrogate matrix ranged from -11.8 to 19.0 %DT with %CV ≤ 14.3. In primary matrix the %DT range was from -5.56 to 5.52, with %CV ≤ 8.67 (**Figure 4B and C**). The accuracy and precision of primary matrix QCs met acceptance criteria and demonstrated the appropriateness and acceptability of using recombinant human MUC5AC and MUC5B as surrogate analytes.

**Figure 4.**
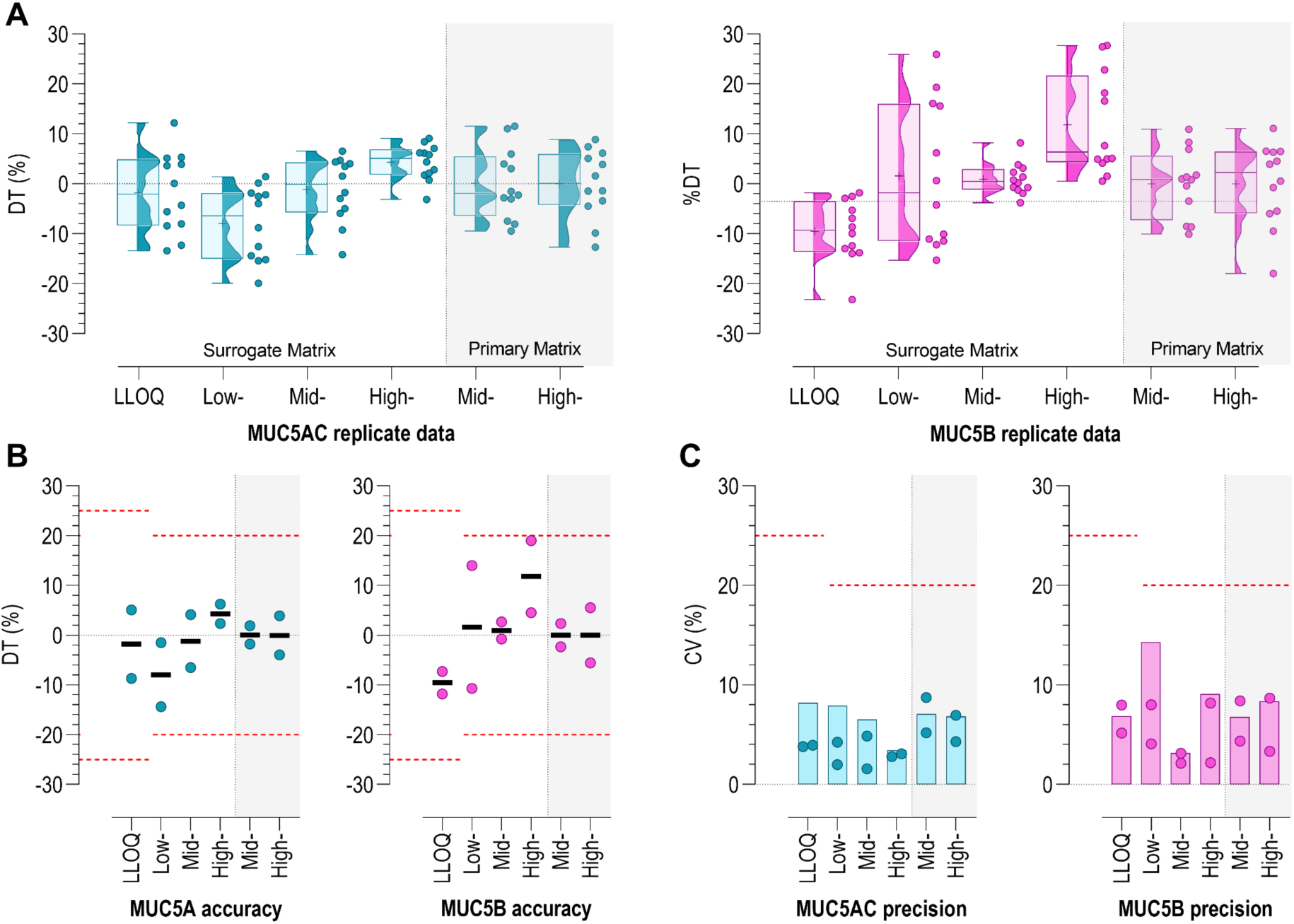
Accuracy and precision for MUC5AC and MUC5B. **(A)** Replicate data for all accuracy and precision runs (n = 2 runs, n = 6 per run). Values are depicted in raincloud plots consisting of jittered univariate dot plots, Tukey-style box-and-whisker plots, and density (half-eye) plots. **(B)** Mean intra- and mean inter-run accuracy values appear as colored dots and as horizontal black lines, respectively. **(C)** Intra- and inter-run precision values appear as colored circles and vertical bars, respectively. Where applicable, acceptance limits are plotted as horizontal, dashed, red lines. LLOQ: lower limit of quantification; QC: quality control; DT: difference from theoretical; CV: coefficient of variation.

#### Specificity and selectivity

Matrix-related background noise and interference can compromise assay performance. Accordingly, six lots of human sputum were evaluated for potential chromatographic interferences at the mass transitions and expected retention times for MUC5AC, MUC5B, and the internal standards. Unfortified specificity samples consisted of double blanks (matrix without IS) and single blanks (matrix with IS) which were extracted and analysed in singlicate or triplicate, respectively. These unfortified specificity samples met acceptance criteria with no appreciable, interfering chromatographic peaks at the retention times of the IS; that is, IS responses in double blanks were ≤ 5% of the mean single blank IS response. Due to the endogenous nature of mucins in human sputum, chromatographic responses were expected for both analytes. As such, formal acceptance criteria were precluded for peaks at the expected retention times of the targeted analytes. Data for the specificity samples were used to establish endogenous concentrations for these six matrices lots and these data were used during the conduct of subsequent parallelism studies.

#### Analyte stability

To evaluate analyte stability in surrogate matrix, under expected general use conditions, we evaluated surrogate matrix QCs at low and high levels. Primary matrix was also used to evaluate general use and handling conditions for unknown samples at endogenous- and overspike-QC levels. For the matrix-related stability experiments (frozen matrix, freeze/thaw, thawed matrix, and overspike), CALs and surrogate matrix-based run acceptance QCs were freshly prepared (never frozen).

To assess analyte stability after multiple freeze/thaw cycles, QCs were subjected to three cycles of thawing (on-ice) and freezing at -80°C. Analyte stability in frozen matrix was evaluated by storing QCs at - 80°C for 42 days prior to analysis. For the thawed matrix analyte stability test, QCs were allowed to thaw and remain on ice for 24h prior to extraction and analysis. To assess, post-preparative extract solution stability, QCs were extracted and injected as part of an analytical run, stored at 2-8°C for approximately 188h, and re-analysed with freshly prepared CALs and QCs.

Analytes were stable in thawed matrix (on-ice ≥24h primary; ≥6.5h surrogate), in post--extraction solutions (≥188h at 2-8°C), and in frozen primary and surrogate matrix (≥188 days at -80°C) (**Figure 5**). For MUC5AC, the analyte was stable in endogenous- and overspike-primary matrix QCs for at least three freeze-thaw cycles (-80°C). For MUC5B, the endogenous-QC demonstrated acceptable analyte stability after three freeze-thaw cycles (-80°C). However, the MUC5B overspike-QC failed to meet acceptance criteria after three freeze-thaw cycles. Accordingly, the overspike-QC evaluation was repeated and found acceptable after two freeze-thaw cycles.

**Figure 5.**
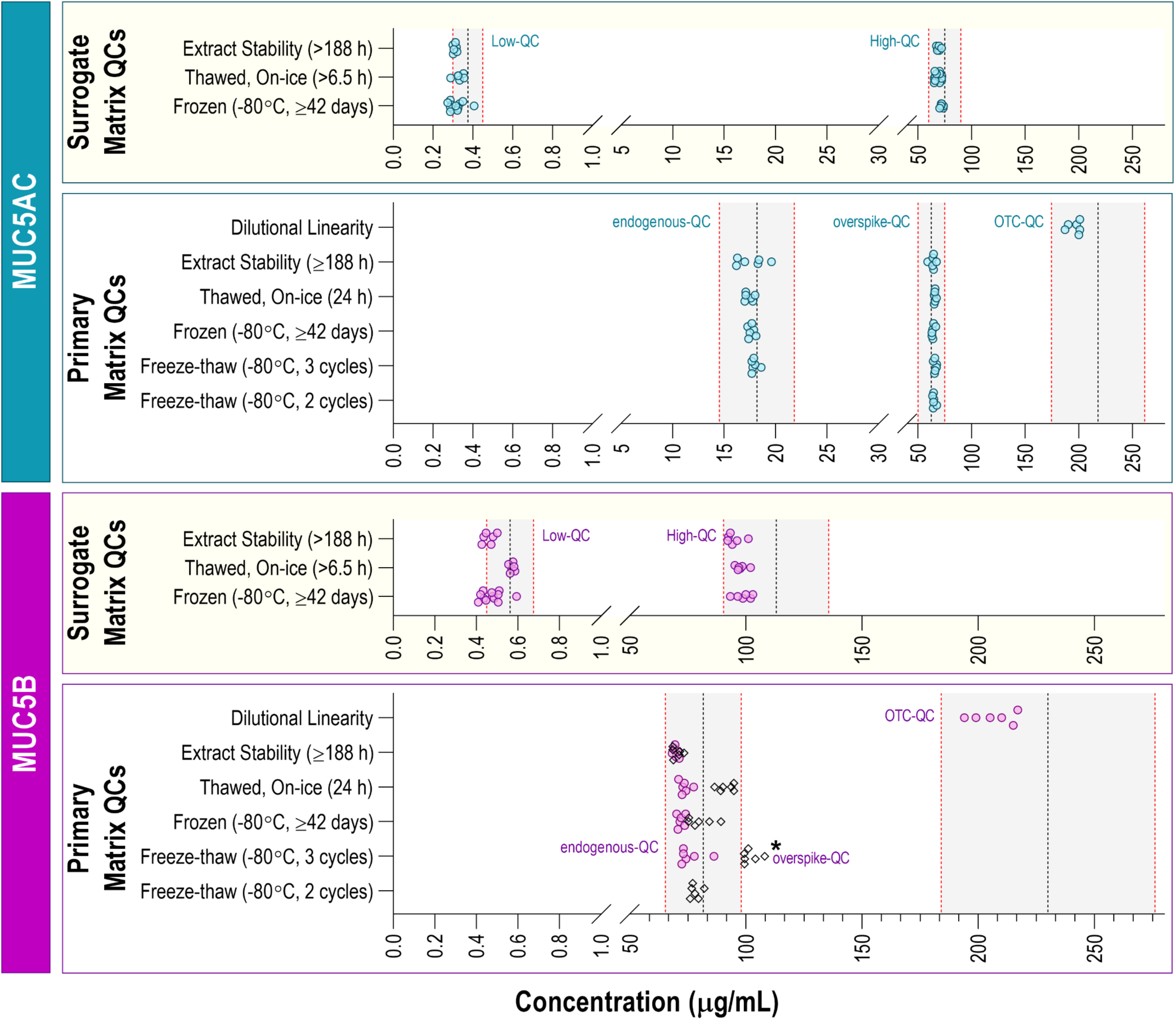
Stability and dilutional study results. Shaded grey bands indicate the acceptance ranges for these studies (±20% from theoretical). At least six replicates were run in each experiment. MUC5B overspike-QCs are represented as black diamonds. The * symbol denotes tests which did not meet acceptance criteria. QC: Quality Control; OTC: Over-the-Curve.

#### Dilutional linearity

To assess the ability to dilute samples with original concentrations well above the calibration range, additional over-the-curve QCs (OTC-QCs) were prepared by spiking pooled primary matrix with rH-MUC5AC or rH-MUC5B such that OTC-QC concentrations for MUC5AC and MUC5B were 218 μg/mL and 230 μg/mL, respectively. These OTC-QCs were diluted 10-fold and analysed in replicate (n = 6). Assay performance was not impacted by diluting samples that were originally above the upper limit of quantification (ULOQ) (**Figure 5**).

#### Matrix parallelism

To demonstrate the appropriateness and acceptability of the surrogate matrix as a stand-in for the primary matrix (matrix parallelism), lots of human sputum (n = 6) were spiked with IS. Mean endogenous MUC5AC and MUC5B concentrations were determined for each IS-spiked matrix lot. Since endogenous analyte concentrations were relatively high, these primary matrix lots were not fortified with additional MUC5AC or MUC5B. Each of the six primary matrix lots were diluted with surrogate matrix and analysed in replicate (n = 3 replicates per dilution level, 2, 8, and 20 fold dilutions) (**Figure 6**). For MUC5AC, all lots of diluted matrix exhibited acceptable precision with %CV ≤ 4.78. For MUC5B, 5 of the six diluted matrix lots exhibited acceptable precision with %CV ≤ 19.76. Accordingly, for this fit-for-purpose method qualification, the surrogate matrix was deemed acceptable and appropriate as a stand-in for the primary matrix.

**Figure 6.**
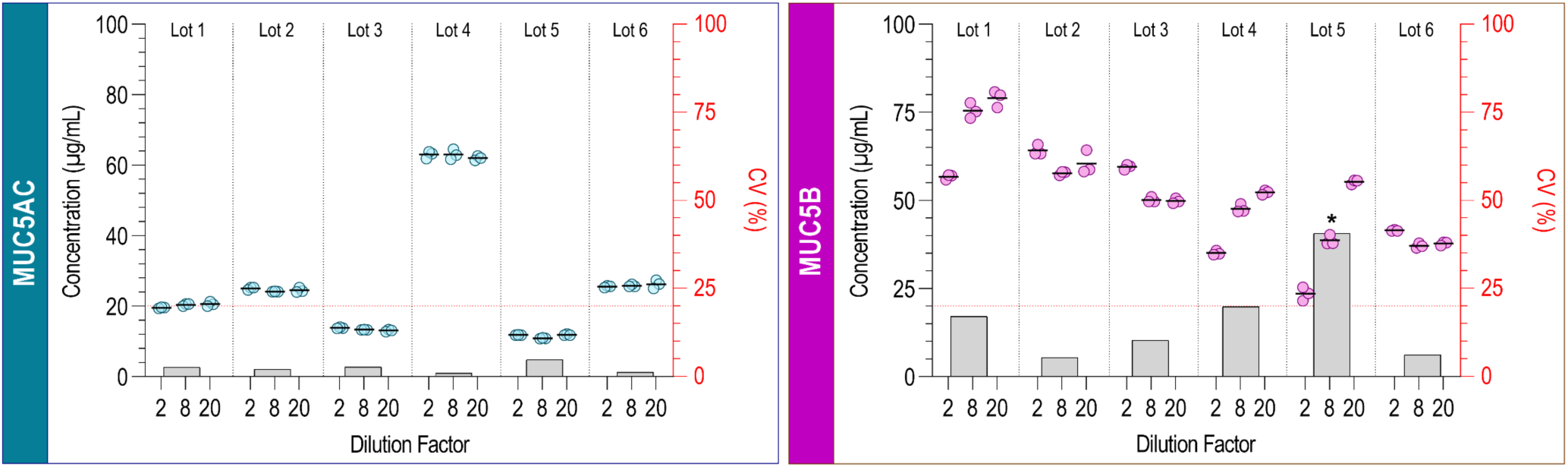
Matrix parallelism. Concentration values for each replicate are depicted as circular data points (n = 3 replicates per dilution, n = 6 lots of human sputum; left axis). Mean intra-dilutional concentrations appear as horizontal, black lines. Mean intra-lot precision values (%CV) appear as vertical bars (right axis). The * symbol denotes matrix lots that did not meet acceptance criteria.

## Conclusions

In this work, we describe a methodology that overcomes many of the challenges related to the heterogenous nature of sputum and the complex physicochemical properties of mucins. We have successfully developed and qualified an LC-MS/MS method for the quantification of soluble MUC5AC and MUC5B in spontaneous human sputum samples using a surrogate analyte and surrogate matrix approach. This analytical method was extensively qualified and was deemed acceptable and appropriate for analysis of clinical samples. In addition, we demonstrated the validity and relevance of airway mucin quantification after total peptide content normalization from frozen spontaneous sputum, which offers added practicality and logistical convenience in a clinical setting. Altogether, our methodology has significant potential to advance the use of mucins as pharmacodynamic biomarkers and aid the development of novel therapeutic strategies targeting mucus hyper-secretion.

## Author Contributions

A.I.R., S.M., K.C., M.G. and I.C.S conceptualization

W.S., K.C., A.I.R, S.M., C.H., H.K., N.A., J.H.L., K.S. and M.Y. methodology

C.H., A.K., H.K. and N.A. resources

W.S., D.V. and S.M. investigation

W.S., K.C., D.V., K.S. and M.Y. formal analysis

D.V., K.S., M.Y., W.W. and A.W. validation

K.C., A.I.R and W.R.M. supervision

A.I.R and K.C. project administration

W.S., K.C., A.I.R., K.S., I.C.S. and C.H. writing - original draft

W.S., K.C., A.I.R., I.C.S., C.H., K.S., M.Y., A.K., S.M., M.G., J.H.L., H.K., N.A. W.W., A.W., W.R.M. and D.V.

writing - review & editing.

## Acknowledgements

The authors thank M. Shane Woolf, PhD (PPD, a part of Thermo Fisher Scientific) for his editorial support and for his assistance with data visualization. We would also like to thank Medicines Evaluation Unit (MEU) for clinical expertise in collecting sputum samples and Sonya Jackson for the development of clinical study plans.

## Writing disclosure

In accordance with current Good Publication Practice (GPP) guidelines, the authors disclose that M. Shane Woolf, PhD (PPD, a part of Thermo Fisher Scientific) provided writing assistance for this manuscript.

## Ethical conduct of research

This study was conducted in accordance with principles of the Declaration of Helsinki and International Conference on Harmonisation Guidance for Good Clinical Practice. Independent ethics committee approval was obtained.

## Data sharing statement

All relevant data required to replicate the study’s findings are within the paper and the supplemental information. Raw data can be obtained in accordance with AstraZeneca’s data sharing policy described at https://astrazenecagrouptrials.pharmacm.com/ST/Submission/Disclosure.

## Supplemental data

This article contains supplemental data.

