## supplemental file for "Development and qualification of an LC-MS/MS method for the quantification of MUC5AC and MUC5B mucins in spontaneous human sputum"

**Supplemental Figure 1. Schematic representation of MUC5AC and MUC5B structures and location of selected quantitative signature peptides.**

**Supplemental Figure 2. Liquid chromatography (LC) mobile phase gradient.**

**Supplemental Figure 3. Representative eight-point calibration curves.**

**Supplemental Figure 4. Representative chromatograms.**

**Supplemental Table 1. Quantitative signature peptides and corresponding internal standards.**

**Supplemental Table 2. MS Instrument parameters.**

**Supplemental Table 3. Demographic information of COPD patients**

### Supplemental Figures

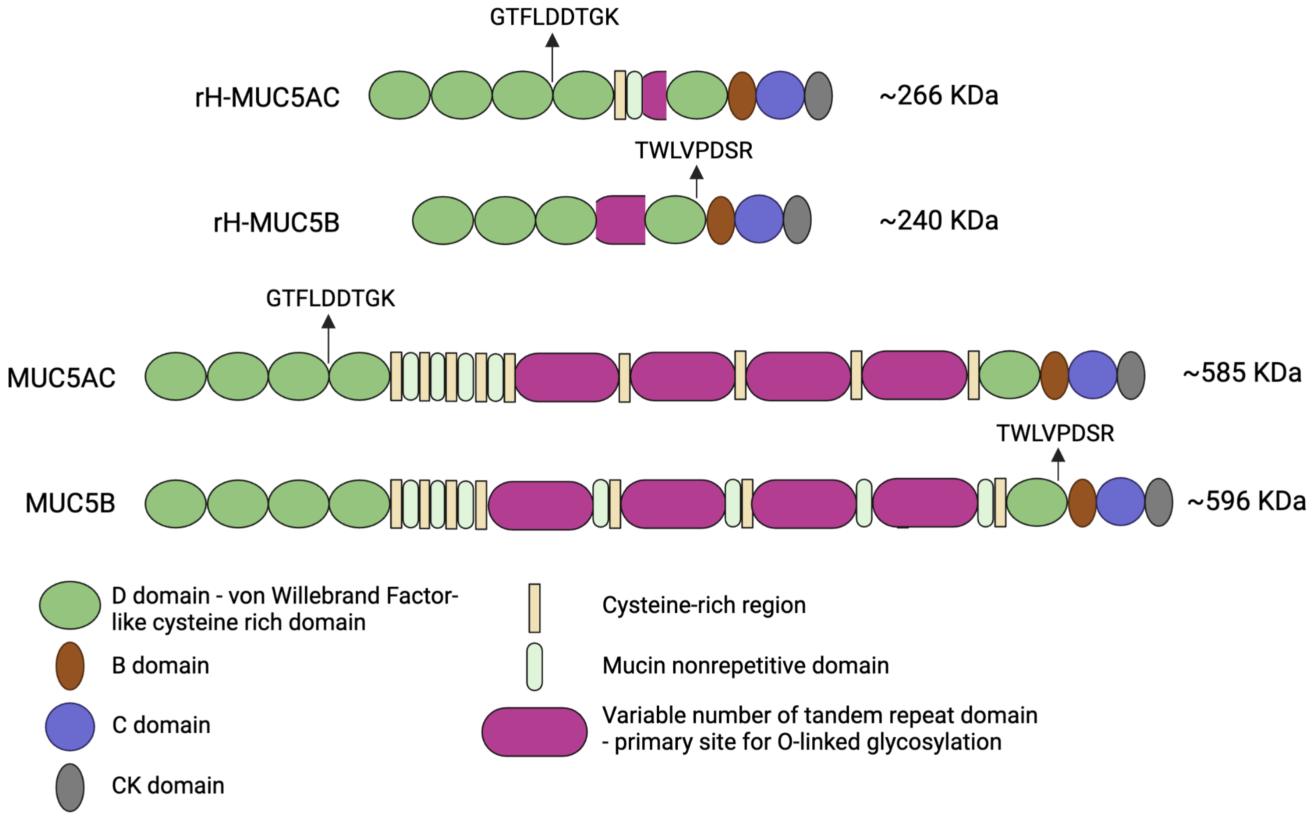


**Supplemental Figure 1. Schematic representation of MUC5AC and MUC5B structures and location of selected quantitative signature peptides.** The rH-MUC5AC construct consists of the N-terminus domain, one tandem repeat, and the C-terminus of MUC5AC, and the rH-MUC5B construct consists of the N-terminus domain, two tandem repeats, and the C-terminus of MUC5B. The calculated molecular weight of unglycosylated proteins is shown. Adapted from Ridley 2018 [1].

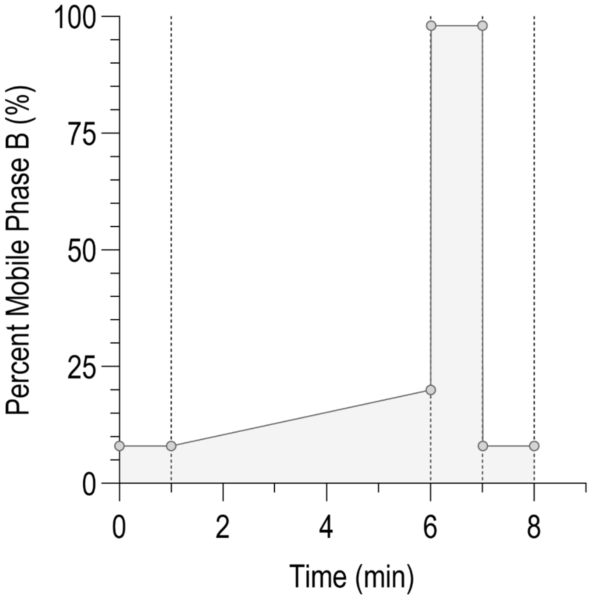


**Supplemental Figure 2. Liquid chromatography (LC) mobile phase gradient.** The LC gradient was started at a composition of 8% mobile phase B for the first 1 minute. Over the next five minutes, the composition of mobile phase B was ramped to 20%. Once the analytes eluted, the column was flushed by increasing the composition of mobile phase B to 98% at 6.01 min, and held at this composition for a minute before returning to the starting %B mobile phase composition of 8% at 7.01 min. The column was allowed to re-equilibrate for an additional minute before the next sample is injected.

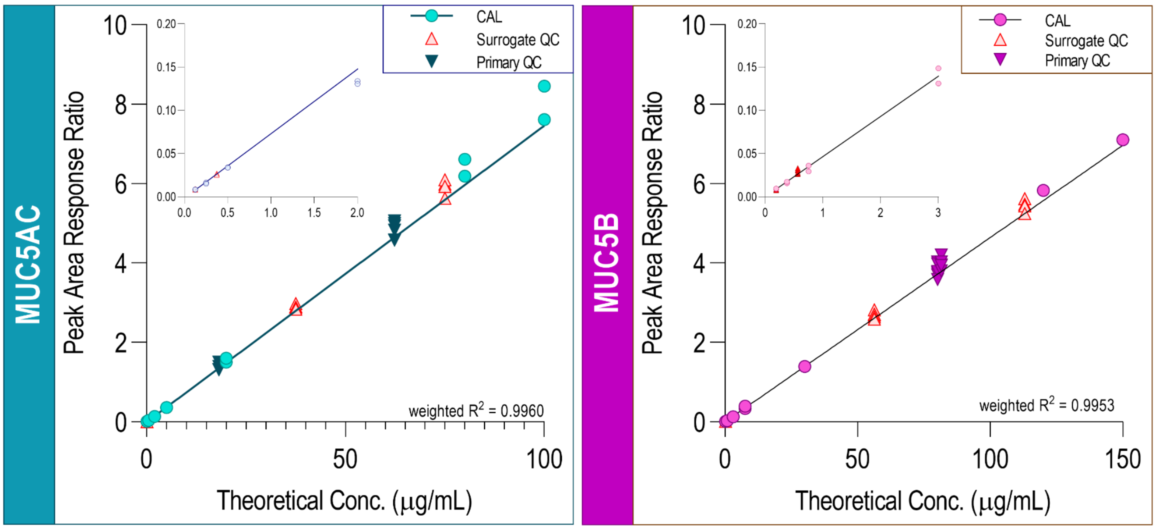


**Supplemental Figure 3. Representative eight-point calibration curves.** Each curve is from a single analytical run with n = 2 replicates per calibrator (CAL) level. Instrument responses for surrogate and primary matrix QCs are plotted against the calibration curves. Note: Primary matrix QCs were diluted; as such, instrument responses for the primary matrix QCs were adjusted to account for this 2-fold dilution. Least squares linear regression: y = mx + b, weighted (1/conc$^2$)

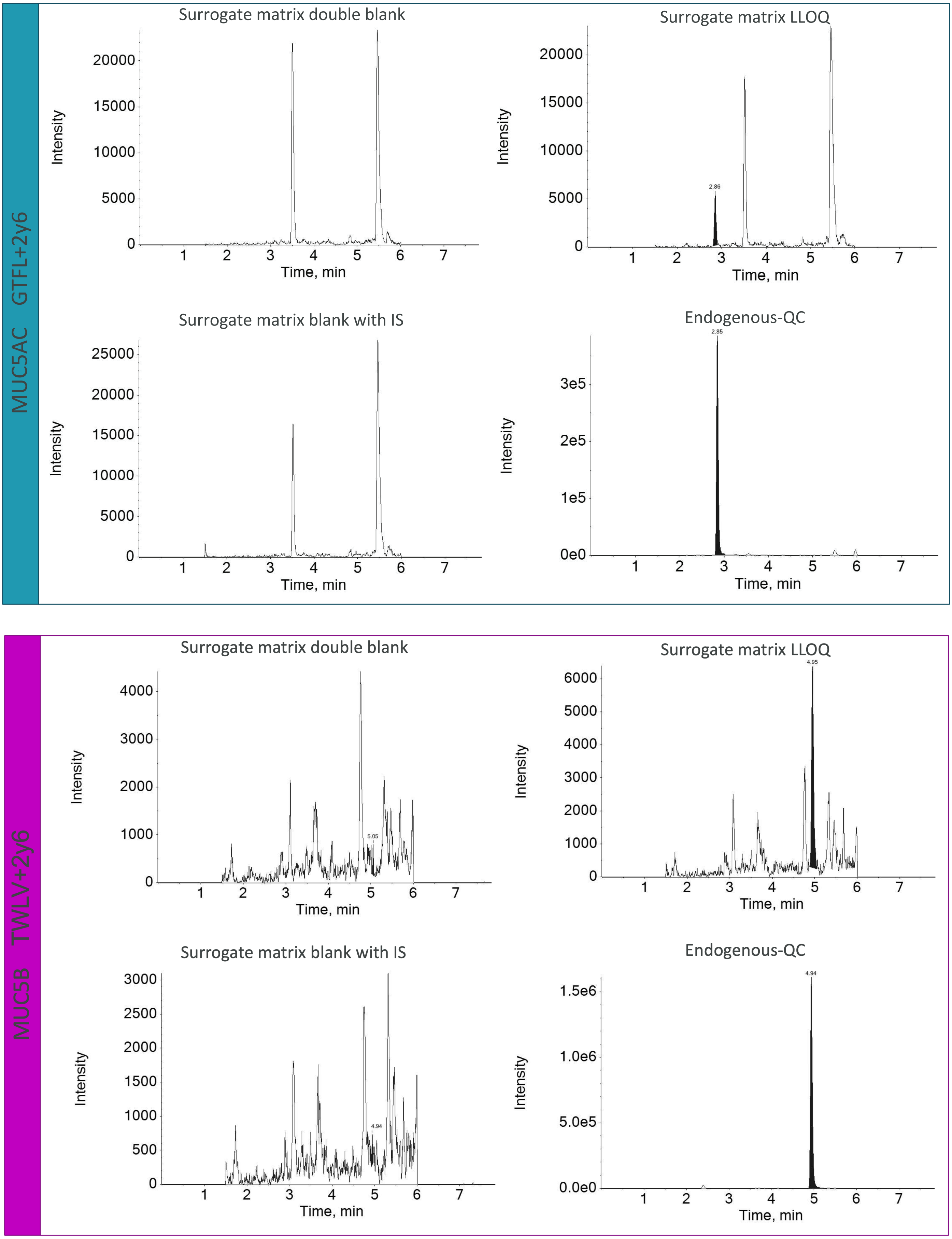


**Supplemental Figure 4. Representative chromatograms.** Integrated peaks are filled with black. Note: Endogenous-QC was diluted by 2-fold.

### Supplemental tables

**Supplemental Table 1. Quantitative signature peptides and corresponding internal standards.**

| Name | Peptide Sequence | Purpose |
|---|---|---|
| GTFL | GTFLDDTGK | Quantitative signature peptide for MUC5AC |
| GTFL-IS | GTFLDDTGK* | Quantitative signature peptide for MUC5AC-IS |
| TWLV | TWLVPDSR | Quantitative signature peptide for MUC5B |
| TWLV-IS | TWLVPDSR* | Quantitative signature peptide for MUC5B-IS |

K* Lys($^{13}C_6$, $^{15}N_2$)
R* Arg($^{13}C_6$, $^{15}N_4$)

**Supplemental Table 2. MS Instrument parameters.**

| Analyte | ~$t_R$ (min) | Q1 m/z | Q3 m/z | Dwell Time (ms) | DP | EP | CE | CXP |
|---|---|---|---|---|---|---|---|---|
| MUC5AC (i.e. GTFL+2y6, Quantitative MUC5AC) | 3.03 | 477.4 | 648.4 | 35 | 32 | 10 | 24 | 20 |
| GTFL-IS (i.e. GTFL+2y6 IS, Quantitative MUC5AC) | 3.03 | 481.4 | 656.4 | 35 | 32 | 10 | 24 | 20 |
| MUC5B (i.e. TWLV+2y6, Quantitative MUC5B) | 5.14 | 487.4 | 686.5 | 35 | 33 | 10 | 22 | 22 |
| TWLV-IS (i.e. TWLV+2y6 IS, Quantitative MUC5AC) | 5.14 | 492.4 | 696.5 | 35 | 33 | 10 | 22 | 22 |

**Supplemental Table 3. Demographic information of COPD patients**

| Diagnosis | Age | Weight (kg) | Height (m) | BMI | Race | Gender | Smoking status |
|---|---|---|---|---|---|---|---|
| **COPD** | 76-80 | 98.7 | 1.70 | 34.0 | Caucasian | Male | Current smoker |
| **COPD** | 71-75 | 68.2 | 1.63 | 25.6 | Caucasian | Female | Ex smoker |
| **COPD** | 71-75 | 79.1 | 1.68 | 27.9 | Caucasian | Male | Ex smoker |
| **COPD** | 61-65 | 49.0 | 1.55 | 20.4 | Caucasian | Female | Current smoker |
| **COPD** | 76-80 | 80.5 | 1.65 | 29.6 | Caucasian | Male | Current smoker |

[1] Ridley C, Thornton DJ. Mucins: the frontline defence of the lung. Biochem Soc Trans. 2018 Oct 19;46(5):1099-1106.
